# Novel Approach to Advance Directive Training: Palliative Fellow Led Workshop Feasible and Effective in Increasing Confidence in End of Life Conversations

**DOI:** 10.1101/2024.03.05.24303812

**Authors:** Chang Chelsea, Jose Cano, Juan Lopez-Alvarenga, Josenny Rodriguez-Paez, Sonya Montes, Meghana Rao

**Affiliations:** Associate Professor Internal Medicine, Division of Primary and Preventive Care, School of Medicine University of Texas Rio Grande Valley, McAllen, Texas, United States of America; Assistant Professor Family Medicine, Division of Primary and Preventive Care, School of Medicine University of Texas Rio Grande Valley, McAllen, Texas, United States of America; Associate Professor of Population Health and Biostatistics, Division of Primary and Preventive Care, School of Medicine University of Texas Rio Grande Valley, McAllen, Texas, United States of America; Internal Medicine Resident, Division of Primary and Preventive Care, University of Texas Rio Grande Valley, School of Medicine, McAllen, Texas, United States of America; Clinical Associate Professor of Medicine Palliative Care, Stony Brook University Hospital. Stony Brook, New York, United States of America

**Keywords:** Advance Directives, Advance Care Planning, Palliative Care, End-of-life, Education

## Abstract

Despite the benefits of Advance directives, approximately only 1 in 3 U.S adults have documented advance directives. In medical school and residency, learners are often not taught or given very brief information on conducting end-of-life planning conversations with patients. Due to this deficiency, some institutions have conducted advance directive workshops but not many have been both palliative fellow and resident led, though some have been led by a geriatric fellow. Therefore, we approached advance directives with a resident and palliative fellow-led workshop. We aimed to develop and conduct a workshop on advance directives to assess feasibility and effectiveness. We sent a survey to 52 residents prior to two one-hour didactic sessions. For the first session, a small group of residents discussed common terminology and the tools available to help patients complete advanced directives in the outpatient setting. A hospice and palliative care fellow led the second session and focused on patient communication and approach. Our results showed that the workshop was well-received and improved resident confidence in discussing advance directives with patients. In conclusion, a resident and palliative fellow-led advance directive workshop for internal medicine residents was feasible and effective in increasing resident confidence.

## Introduction

Despite the benefits of Advance directives, approximately only 1 in 3 of U.S adults have documented advance directives [1-3]. In medical school and residency, learners are often not taught or given very brief information on how to conduct end-of-life planning conversations with patients [4-6]. Due to this deficiency, some institutions have conducted advance directive workshops, but not many have been both resident and palliative care fellow-led, though some have been led by a geriatric fellow [3,7]. Hospice and Palliative fellows have increased exposure to encounters related to end-of-life conversations compared to other residents and therefore are an excellent resource for this needed education[8,9]. We decided to approach advanced directives with a resident and palliative fellow-led workshop.

Advance care planning is a way to help patients and families make important decisions in planning for their future. It is crucial in end-of-life care and can significantly ease the emotional burden in critical situations. Discussing advance directives early on has been shown to have better outcomes in families [10-12]. Lack of advance directive documentation is compounded in the Rio Grande Valley of South Texas, where patient health literacy rates are low, and discussions regarding end of life are generally avoided in the Hispanic culture, making these conversations even more challenging [13,14]. These pose barriers to helping patients plan for their future.

Palliative care fellows must demonstrate competence in coordinating, leading, and facilitating advance directive completion. Residents and fellows must also complete a scholarly or quality improvement project during their programs; therefore, this met educational and patient needs. We reviewed and implemented proven strategies to change physician behavior and to teach communication skills to physicians to include demonstration and role-play[15-18]. We aimed to develop and conduct a resident and palliative fellow-led workshop on advance directives to assess feasibility and effectiveness.

## Methods

We sent a survey to 52 Internal Medicine Residents prior to two one-hour didactic sessions. Our team presented two one-hour workshops, one resident-led and one palliative fellow-led, aiming to raise resident confidence in discussing advance directives. A small group of residents discussed common terminology and the tools available to help patients complete advance directives in the outpatient setting in their lecture hall using a PowerPoint presentation. The resident presenters discussed the four types of advance directives including medical power of attorney, reviewed the Texas advance directive document and discussed ways to make the document official. A hospice and palliative care fellow led the second session and focused on patient communication and approach. The fellow-led workshop included the use of prognostication tools such as the Palliative performance scale, FAST, and ECOG. The fellow presented a PowerPoint including advance directive terminology, prognostication tools, and a bar graph representation of pre-survey results. The palliative fellow then demonstrated an example of an advance directive conversation with the help of a resident volunteer, who played the role of a patient. The palliative fellow then instructed the residents to pair up and role-play two provided patient scenarios. Instructions included that they had ten minutes to role-play each scenario, with one resident as a physician and the other as a patient. They subsequently switched roles for the second scenario. We provided the residents with prognostication tool handouts and printed scenarios. The first case was NYHA Class IV heart failure, and the second was a case of metastatic cancer. The palliative fellow then conducted a two-minute debriefing session after each scenario to discuss questions or comments. The palliative fellow then discussed a third Alzheimer’s dementia patient scenario with the group as a whole and followed it up with a post-survey that was identical to the pre-survey.

We used a 5 point Likert agreement scale to assess the residents’ level of confidence pre and post-survey. We conducted a pre and post-test comparison for independent groups using a t-test adjusted for unequal variances.

We hypothesized that the workshop would increase residents’ knowledge and confidence in advance care planning discussions. Ethical aspects include that UTRGV IRB determined this project to be exempt.

## Results

Regarding feasibility, with two faculty mentors, the group of internal medicine residents and fellow were able to develop and lead the two sessions within one academic year. A total of 52 Internal Medicine Residents were invited, 42 (80%) completed the pre survey and 34 (65%) completed the post survey.

The 5 questions with pre and post means are in Table 1 with the Likert Scale of 1 being strongly disagree and 5 strongly agree Table 1. Q1 [I have sufficient knowledge of advance directives, given my years of training] showed a pre-test mean of 3.0 and a post-test mean of 3.8, p-value 0.024. Q2 [I believe my experience with advance directives is adequate for the situations that I routinely encounter] showed a pre-test mean of 3.0 and a post-test mean of 3.9, p-value of <0.001.

**Table 1.**
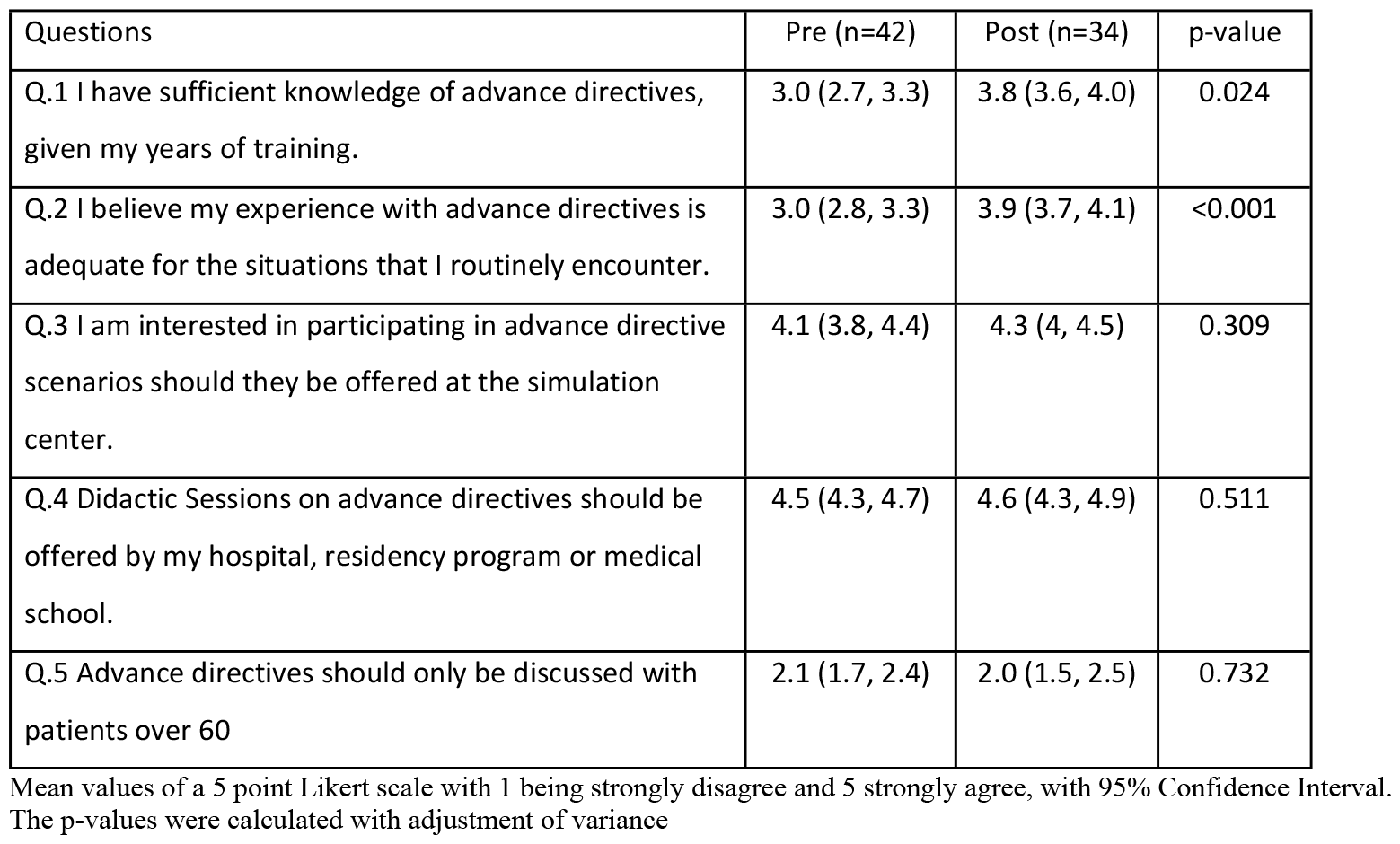
Pre and Post Survey of Advance Directive Training.

| Questions | Pre (n=42) | Post (n=34) | p-value |
| --- | --- | --- | --- |
| Q.1 I have sufficient knowledge of advance directives, given my years of training. | 3.0 (2.7, 3.3) | 3.8 (3.6, 4.0) | 0.024 |
| Q.2 I believe my experience with advance directives is adequate for the situations that I routinely encounter. | 3.0 (2.8, 3.3) | 3.9 (3.7, 4.1) | <0.001 |
| Q.3 I am interested in participating in advance directive scenarios should they be offered at the simulation center. | 4.1 (3.8, 4.4) | 4.3 (4, 4.5) | 0.309 |
| Q.4 Didactic Sessions on advance directives should be offered by my hospital, residency program or medical school. | 4.5 (4.3, 4.7) | 4.6 (4.3, 4.9) | 0.511 |
| Q.5 Advance directives should only be discussed with patients over 60 | 2.1 (1.7, 2.4) | 2.0 (1.5, 2.5) | 0.732 |

Mean values of a 5 point Likert scale with 1 being strongly disagree and 5 strongly agree, with 95% Confidence Interval. The p-values were calculated with adjustment of variance

Both perceived knowledge (Q1) and confidence (Q2) had a statistically significant move from a response of “neutral” (3) to a response of “agree” (4) and are depicted in histograms to show the distribution of responses improving Figure 1 and 2.

**Fig 1.**
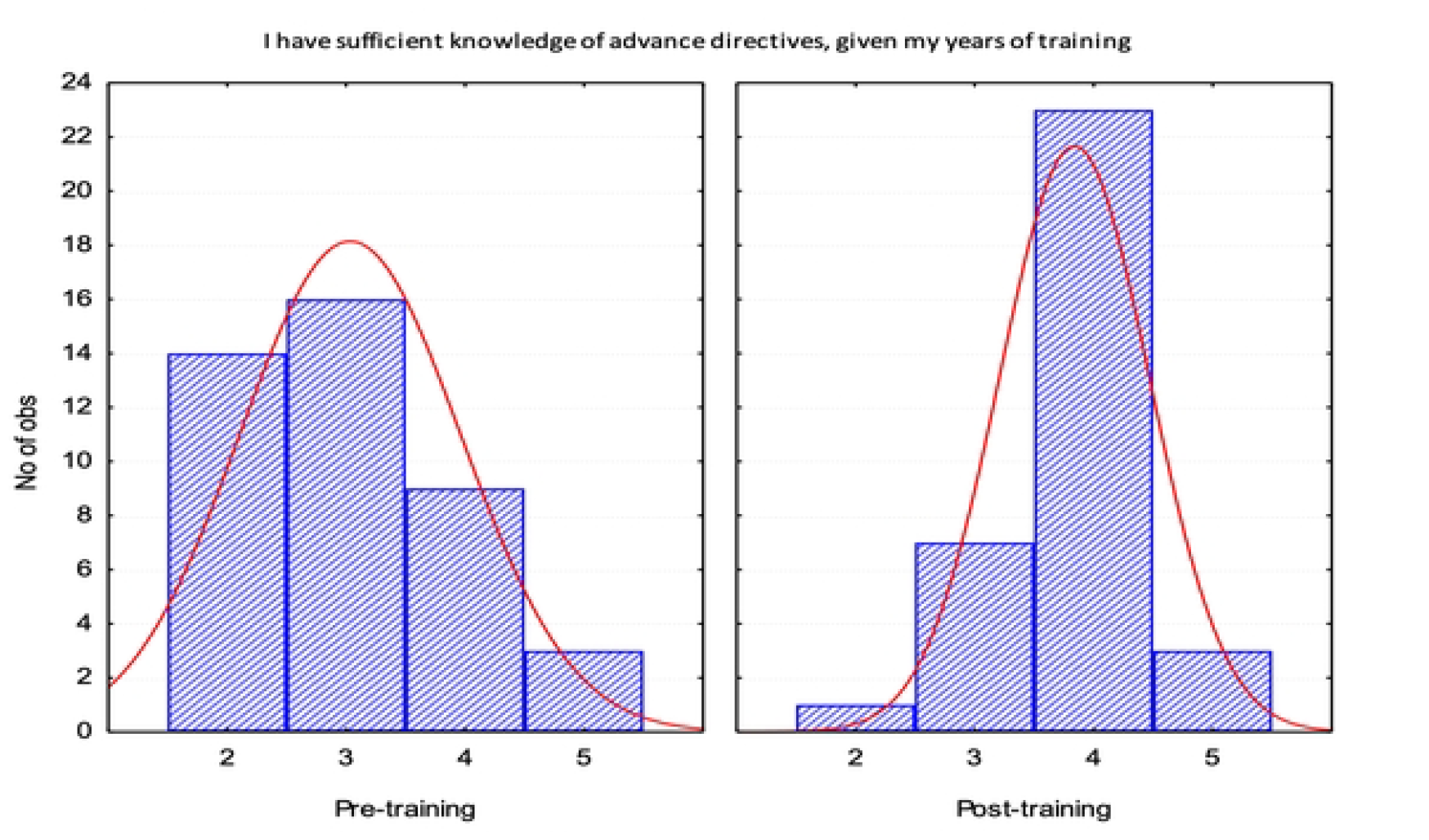
Histogram Post Training Survey. X axis 5-point Likert scale with 1 being strongly disagree and 5 strongly agree. Y axis represents N of observations or participants. Right Curve Deviation on Post Test representing improvement (Observe in the post-training distribution a clear displacement of the curve to the right on the x-axis).

**Fig 2.**
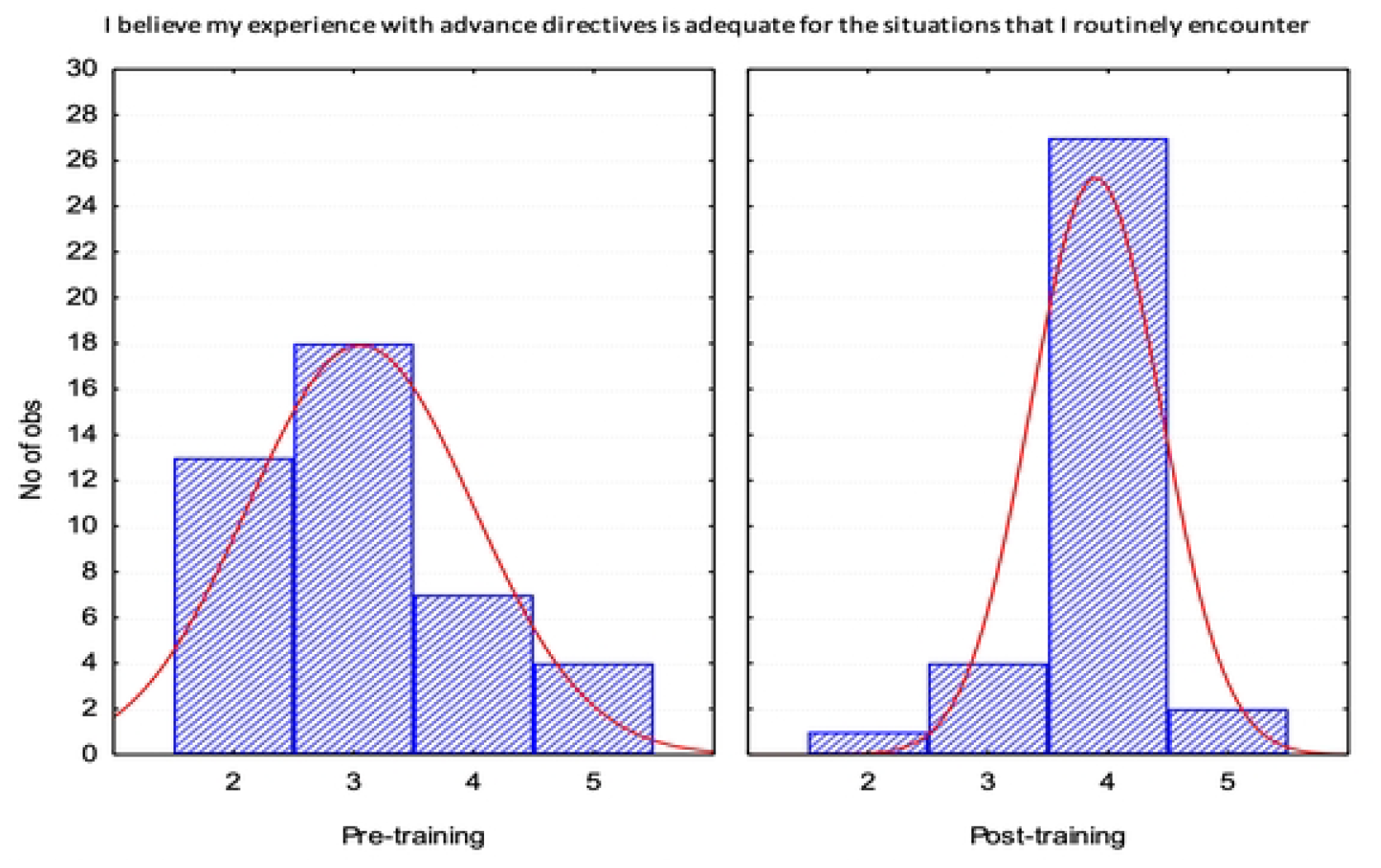
Histogram Post Training Survey. X axis 5-point Likert scale with 1 being strongly disagree and 5 strongly agree. Y axis represents N of observations or participants. Right Curve Deviation on Post Test representing improvement (Observe in the post-training distribution a clear displacement of the curve to the right on the x-axis).

Resident interest in participating in advance directive simulations and resident agreement that didactics on advance directives should be offered, both had high agreement pre and post with no statistically significant change (Q3 and Q4 in Table 1). Q5 assessed knowledge directly with the statement that advance directives should only be discussed with patients over 60 to which both pre and post means were correct in “disagreeing”, with no statistically significant change.

During the workshops, residents expressed that practicing patient scenarios gave them insight into navigating these types of conversations and found this method impactful. Residents also verbalized that exposure to prognostication tools helped in these discussions as well. Residents reported that experiencing both physician and patient roles also exposed them to the barriers that each position entails in advance directive conversations.

Thus, overall, our results showed that the workshop was well-received and improved resident confidence in discussing advance directives with patients.

## Discussion

Our fellow and resident-led advance directive workshop was feasible as evidenced by completion and effective as evidenced by pre and post-surveys demonstrating statistically significant increased knowledge and confidence. The observed high level of resident interest in training, by simulation or didactics, in both pre and post survey also support the notion that these workshops are needed. Upon reflection of our knowledge-based question regarding whether to only perform directives in people above 60 years of age, the majority of residents already knew this statement was incorrect in the pretest, therefore no change was accomplished in posttest. This likely indicates the question was too simple and did not assess the higher order of knowledge which we were seeking to impart.

A strength of this project is that this model of resident and fellow-led advance directive workshop fills a curricular gap for internal medicine residents while also satisfying educational requirements for both residents and fellows. The fellow provided a near-peer teaching environment, and resident leaders appreciated that additional level of experience and expertise the fellow added. Another strength is overall high pretest completion at 80% with slightly lower but still above average posttest completion of 65%. Our workshop added to the literature showing success in resident-led workshops, improved confidence in advance directive discussions after training and was unique in the involvement of a palliative care fellow[3,7]. Our model may be a tactic to address the deficiency in end-of-life planning among resident learners at institutions with palliative care fellows and provide a successful method in bolstering resident confidence in discussing advance directives.

Limitations included that survey responses were not matched to individuals which lowered statistical efficiency in detecting individual changes, surveys were not validated and mostly assessed Kirkpatrick Level 1 Reaction with a small amount of Kirkpatrick Level 2 Learning on knowledge.

Regarding feasibility limitations, we performed the workshop at a single site and implemented it in a single year. Future studies can include assessing Level 3 Behavior or Level 4 Results such as in effectiveness and frequency in which residents engage in advance directive discussions.

## Conclusion

Advance care planning plays a pivotal role in end-of-life care. Through our two-session workshop, we improved resident confidence in discussing end-of-life planning. In conclusion, a resident and palliative fellow-led advance directive workshop for internal medicine residents was feasible and effective in increasing resident confidence.

## Data Availability

All Relevant data are within the manuscript and its Supporting Information files.

## Supporting information

S1 Fig,1. **Fig 1. Histogram Post Training Survey**. X axis 5-point Likert scale with 1 being strongly disagree and 5 strongly agree. Y axis represents N of observations or participants. Right Curve Deviation on Post Test representing improvement (Observe in the post-training distribution a clear displacement of the curve to the right on the x-axis).

**S2 Fig 2. Histogram Post Training Survey**. X axis 5-point Likert scale with 1 being strongly disagree and 5 strongly agree. Y axis represents N of observations or participants. Right Curve Deviation on Post Test representing improvement (Observe in the post-training distribution a clear displacement of the curve to the right on the x-axis).

